# Prognostic Value of Patient-Reported Outcomes in Predicting Long-term Mortality after Transcatheter Aortic Valve Replacement (TAVR)

**DOI:** 10.1101/2023.03.29.23287935

**Authors:** Hasan S. Alarouri, Alejandra Chavez Ponce, Abdulah Mahayni, Sahar Samimi, Mohamad Alkhouli

**Author notes:** **Corresponding Author:** Mohamad Alkhouli, MD Professor of Medicine, Mayo Clinic | 200 First Street SW | Rochester, MN 55905. **Funding:** No funding was obtained for this research.

## Abstract

**Background:** Patient-reported outcome measures (PROM) have been shown to have important prognostic value after various cardiac interventions. We assessed the association between the change in Kansas City Cardiomyopathy Questionnaire 12 (KCCQ-12) score after transcatheter aortic valve replacement (TAVR) and mortality.

**Methods:** We included patients who underwent TAVR at Mayo Clinic between February 2012, and June 2022, who completed KCCQ-12 prior to and 30-45 days after the procedure. Patients were categorized into 3 groups: those who experienced significant (≥ 19 points; **group-1**), modest (1-19 points; **group-2**), and no improvement (≤ 0 points; **group-3**).

**Results:** A total of 1,124 patients were included: 60.8% males; 97.6% Caucasian. Mean age was 79.4±8.3 years, KCCQ-12 score was 53.9±24.5, and median STS score was 4.9% (interquartile range 3.1-8.0). At 45 days, the mean change in KCCQ-12 score was 19±24 points: 46.3% (n= 520) of patients had a significant improvement in their KCCQ-12 score, while 33.4% (n= 375) and 20.4% (n = 229) had modest and no improvement, respectively. Median survival was higher in group-1 (5.7±0.2 years) compared to groups 2 and 3 (5.1±0.3 and 4.1±0.4 years, respectively; *P*<.001). Compared to patients in group 1, those in groups 2 and 3 had higher long-term risk-adjusted mortality (adjusted hazard ratio 1.64; 95%CI 1.28-2.10, and 2.44; 95%CI, 1.84-3.24, respectively).

**Conclusion:** Patients who experience modest or no improvement in KCCQ-12 score after TAVR have substantially higher long-term mortality. Delta KCCQ-12 is a cost-effective, efficient tool that can identify patients at increased risk of death at long-term follow-up post-TAVR.

**Clinical Perspective:** 

**What is New?:**

- This study documents the prognostic value of the Kansas City Cardiomyopathy Questionnaire 12 score in predicting long-term survival after TAVR.
- Modest or no improvement in the Kansas City Cardiomyopathy Questionnaire 12 score after TAVR is associated with a considerably higher long-term mortality risk.

**What are the Clinical Implications?:**

- Changes in the Kansas City Cardiomyopathy Questionnaire 12 score may identify patients with higher residual mortality risk after transcatheter aortic valve replacement.

---

Transcatheter aortic valve replacement (TAVR) has revolutionized the management of severe aortic stenosis (AS) worldwide. With the continued demonstration of excellent outcomes, the indication for TAVR has expanded and the annual procedural volumes have grown exponentially. However, a considerable proportion of patients undergoing TAVR are either dead or experience no symptomatic improvement at 1 year, suggesting the need for additional strategies to identify and mitigate residual risks.^1^

The Kansas City Cardiomyopathy Questionnaire (KCCQ) is a commonly utilized instrument that assesses the quality of life in patients. Originally designed to be used in patients with heart failure (HF), KCCQ has been validated as a prognostic tool in a variety of procedural settings.^2–5^ In additional, the KCCQ score was later abbreviated and extensively validated to allow its scalability and wide usage.^6^ The modified version, the KCCQ-12 has been adopted and validated to assess patients’ quality of life prior to and after the treatment of various cardiac conditions, including AS.^7–9^ Recently, the KCCQ-12 demonstrated prognostic capabilities in predicting 1-year mortality and HF hospitalization among patients undergoing TAVR.^10,11^ However, to our knowledge, there are limited data on the association between delta KCCQ-12 after TAVR and long-term survival. We hence explored this association leveraging prospectively collected data from a large tertiary valve center.

## Methods

### Data Availability

As the study utilized data of a sensitive nature, any requests to access this data must be from qualified researchers who have received adequate training regarding human subject confidentiality protocols. Requests may be submitted to the corresponding author.

### Study Population

We utilized the Society of Thoracic Surgeons (STS) and American college of Cardiology’s (ACC) Transcatheter Valve Therapy (TVT) registry of patients who underwent TAVR at Mayo Clinic Hospital (Rochester, Minnesota) from February 8^th^, 2012, to June 30^th^, 2022. Eligibility for TAVR was determined by a heart-team consisting of an interventional cardiologist and cardiothoracic surgeon and primarily guided by the Food and Drug Association (FDA) and ACC/American Heart Association’s (AHA) eligibility criteria.^12,13^ We included patients who underwent the procedure and who had both a preprocedural and a postprocedural KCCQ-12 administered (n=1,124). The postprocedural KCCQ-12 was administered 30-45 days after the procedure, which we defined as the early follow-up visit. Patients whose vital status was censored within 45 days of the procedure (n=63) and those who did not have both questionnaires administered (n=1,629) were excluded from our study. A comparison of baseline characteristics between patients included in our study and those who were excluded due to unavailable KCCQ-12 data is summarized in **Table S1** of the **Supplement**.

### Study Endpoint and Design

The primary endpoint of our study was the association between delta KCCQ-12 score and long-term mortality in TAVR patients. Secondary outcomes included: association of baseline KCCQ-12 score and long-term mortality, difference in median survival time stratified based on the degree of change in the KCCQ-12 score. We defined a significant improvement in the KCCQ-12 score from baseline as one equal to or greater than the mean change in score at the early follow-up visit (+19 points). Subsequently, we categorized patients into three groups; those who experienced a significant improvement (≥ +19 points; group-1), modest improvement (1-19 points; group-2), and no improvement (≤ 0 points; group-3). This study was approved by the Mayo Clinic Institutional Review Board, and a requirement for patient consent was waived as it was considered a minimal risk, retrospective research study.

### Survey instrument (KCCQ-12)

The KCCQ-12 is a health-status assessment tool that consists of 12 items enclosed within the following four domains: symptoms, functional status, quality of life, and social limitations (**Figure S1**).^6^ It produces a scaled score ranging from 0 (denoting the worst possible score) to 100 (denoting the best possible score) points.

### Data Analysis

Categorical variables were reported as frequencies with percentages, while continuous variables were reported as either mean values with the standard deviation (SD) or median values with interquartile ranges (IQR) for skewed distributions. We compared procedural, in-hospital, and early follow-up outcomes across the 3 groups using either a Pearson’s chi-square or Fisher’s exact test for categorical variables, and a one-way analysis of variance (ANOVA) or Kruskal-Wallis test for continuous variables. Median follow-up time was computed from the date of the procedure to the date of death or censoring. Long-term survival was estimated using the Kaplan-Meier method. Adjusted hazard ratios (HR) were calculated using a Cox-proportional hazards regression model and were depicted with their 95% confidence interval (CI). The model adjusted for potential confounders and group differences in baseline characteristics with a *P* value <0.05; thus, the following covariables were adjusted for: age, atrial fibrillation, AV mean gradient, LVEF, NYHA class, baseline KCCQ-12 score, body mass index, chronic pulmonary disease, diabetes, hemodialysis, hypertension, immunocompromised status, postprocedural pacemaker, TAVR indication, prior myocardial infarction, prior cardiac surgery, prior stroke or transient ischemic attack (TIA), sex, smoking status, and Society of Thoracic Surgeons (STS) risk score. A *P* value <.05 was considered statistically significant. Statistical analyses were conducted using Statistics Package for the Social Sciences (SPSS), version 28.0.0.0, and GraphPad Prism, version 9.3.1.

## Results

### Study Characteristics

Our study examined 1,224 patients (39.2% females; 97.6% Caucasian) who underwent TAVR between February 8^th^, 2012, to June 30^th^, 2022. At the time of the procedure, the overall mean (±SD) age was 79.4±8.3 years, KCCQ-12 score was 53.9±24.5 points, aortic valve mean gradient was 42.7±13.2 mmHg, and aortic valve peak velocity was 4.1±0.7 m/s (**Table 1**). The median STS risk score was 4.9% (IQR, 3.1-8.0), and 67.0% of patients had an NYHA class of III or IV.

**Table 1.**
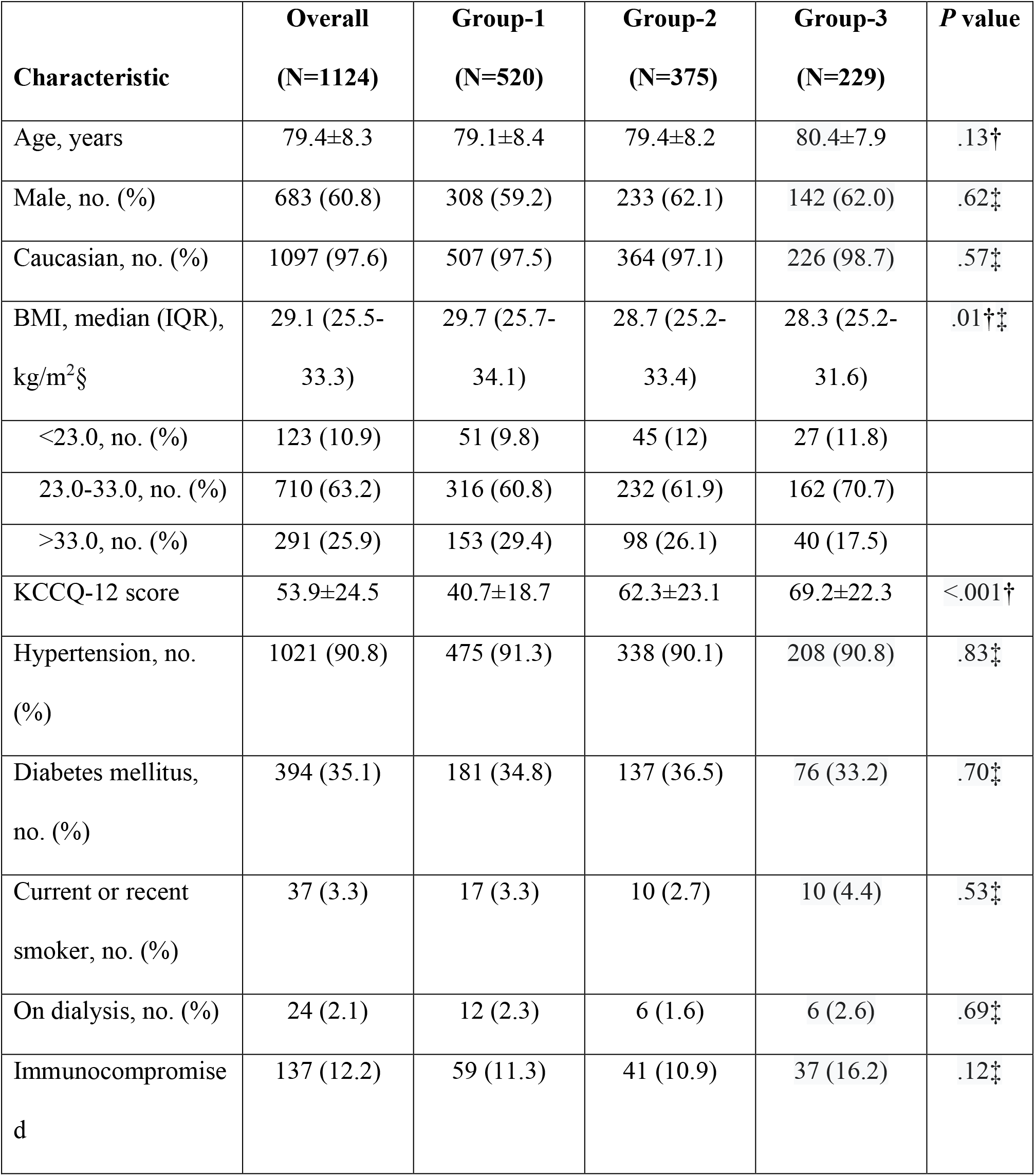
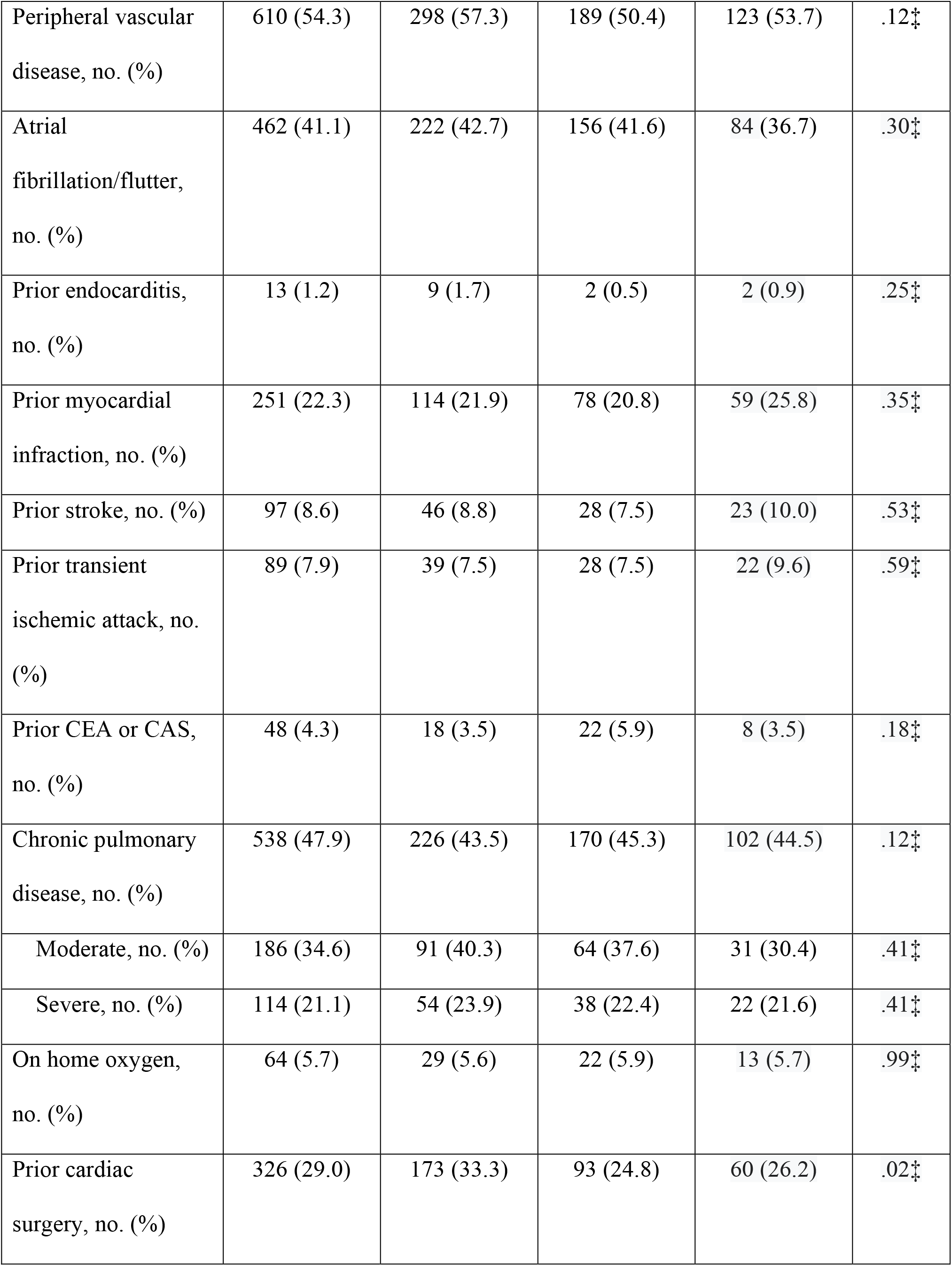
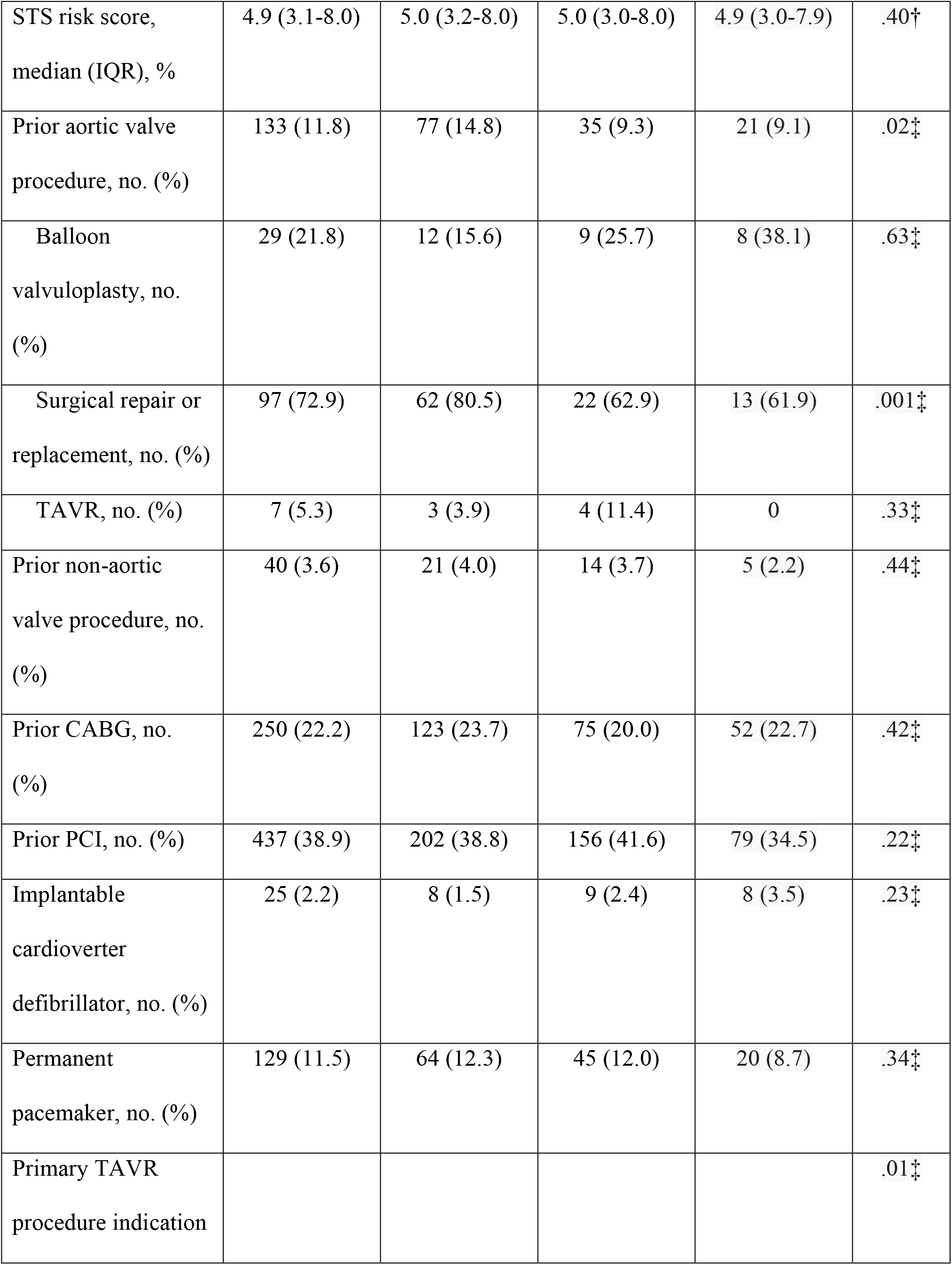
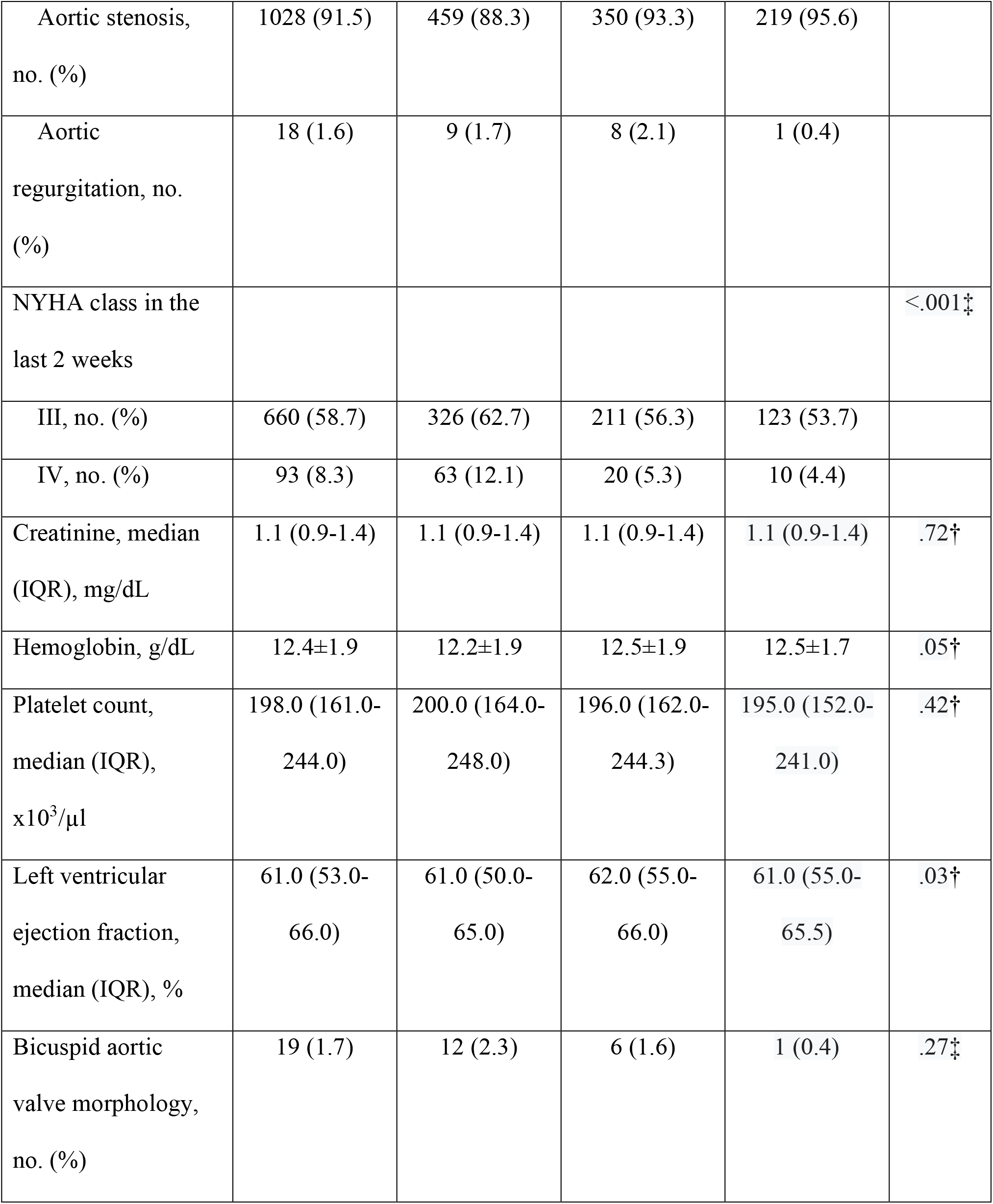
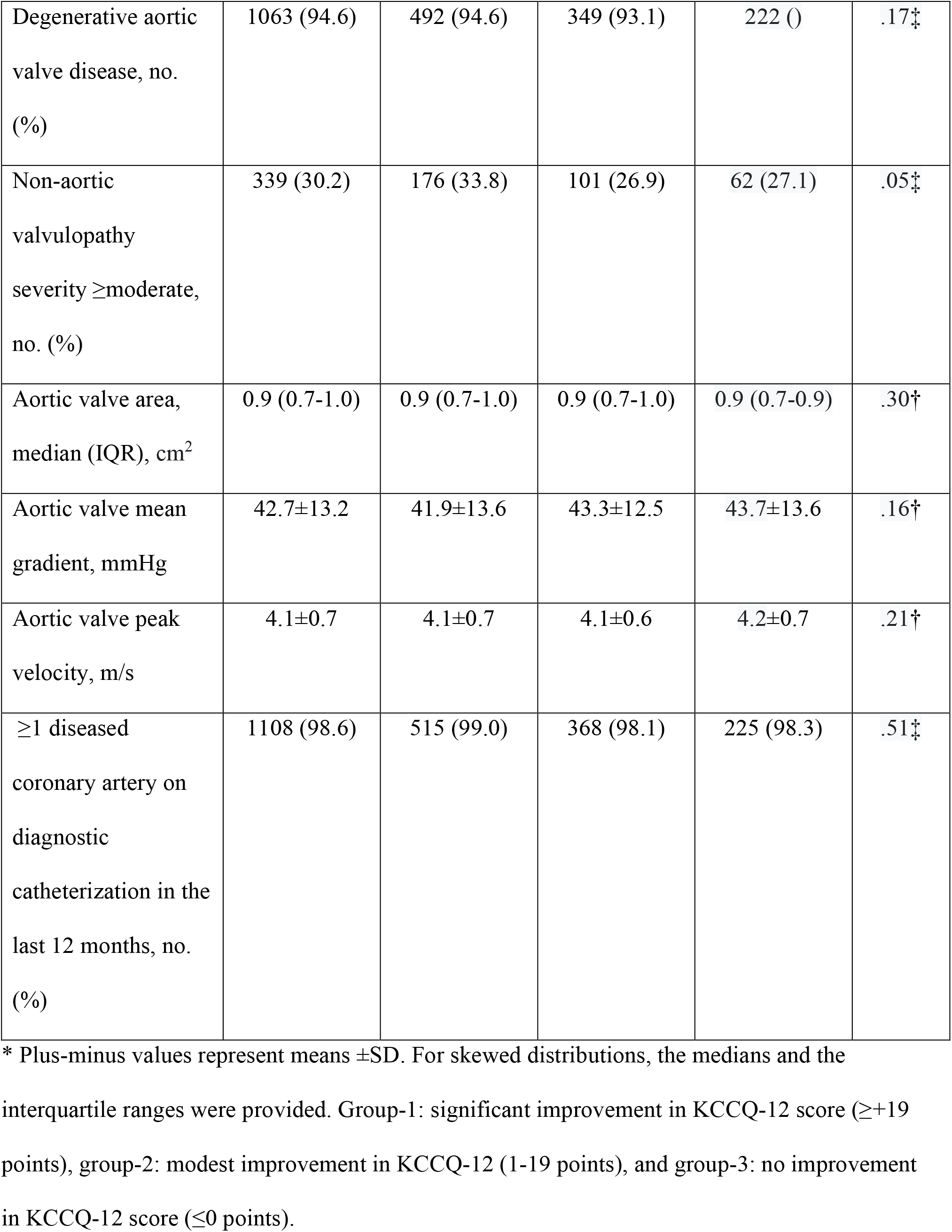
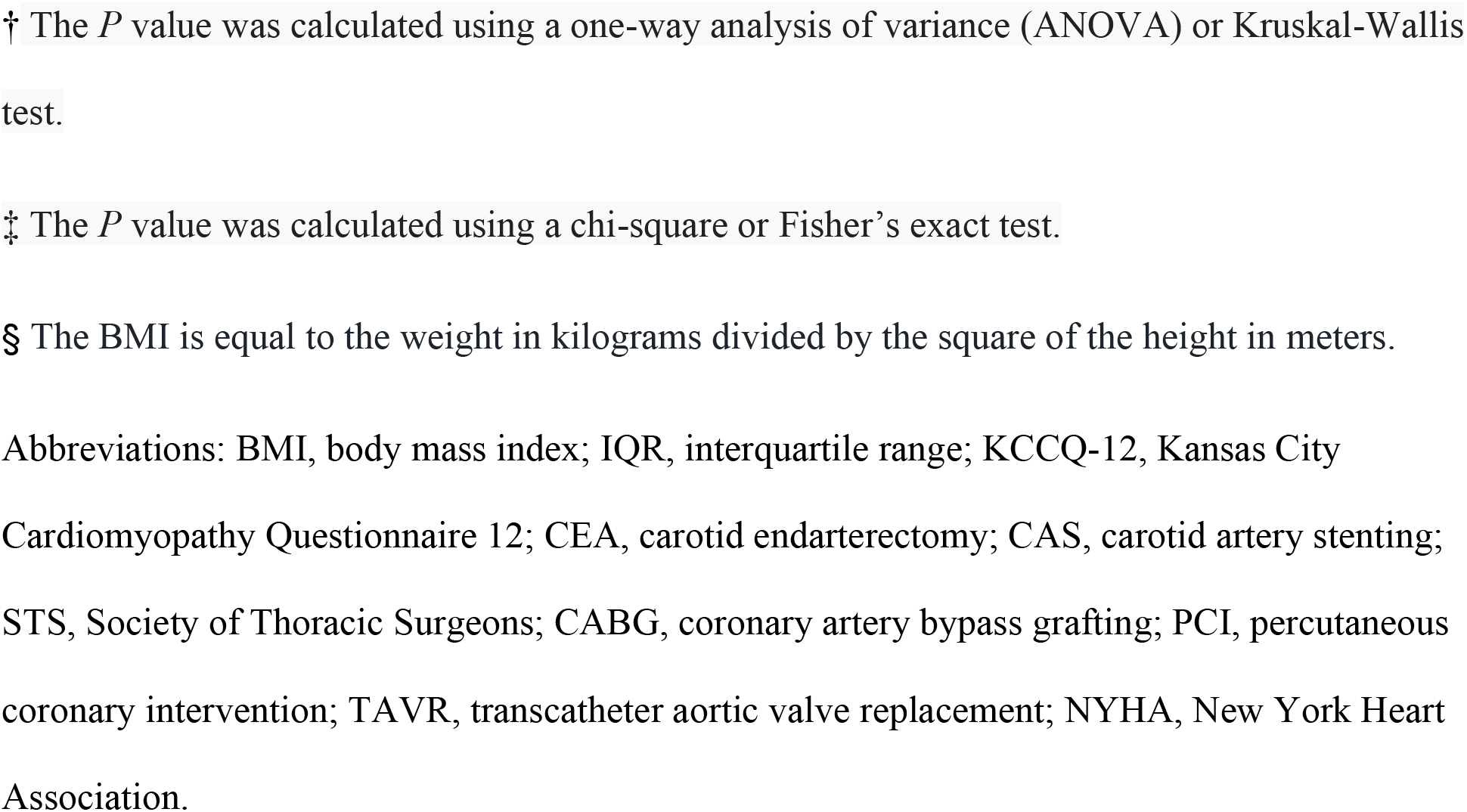
Overall and Comparison of Baseline Clinical and Echocardiographic Characteristics *.

| **Characteristic** | **Overall**<br>**(N=1124)** | **Group-1**<br>**(N=520)** | **Group-2**<br>**(N=375)** | **Group-3**<br>**(N=229)** | **P value** |
| --- | --- | --- | --- | --- | --- |
| Age, years | 79.4±8.3 | 79.1±8.4 | 79.4±8.2 | 80.4±7.9 | .13† |
| Male, no. (%) | 683 (60.8) | 308 (59.2) | 233 (62.1) | 142 (62.0) | .62‡ |
| Caucasian, no. (%) | 1097 (97.6) | 507 (97.5) | 364 (97.1) | 226 (98.7) | .57‡ |
| BMI, median (IQR),<br>kg/m<sup>2</sup>§ | 29.1 (25.5-33.3) | 29.7 (25.7-34.1) | 28.7 (25.2-33.4) | 28.3 (25.2-31.6) | .01†‡ |
| <23.0, no. (%) | 123 (10.9) | 51 (9.8) | 45 (12) | 27 (11.8) |  |
| 23.0-33.0, no. (%) | 710 (63.2) | 316 (60.8) | 232 (61.9) | 162 (70.7) |  |
| >33.0, no. (%) | 291 (25.9) | 153 (29.4) | 98 (26.1) | 40 (17.5) |  |
| KCCQ-12 score | 53.9±24.5 | 40.7±18.7 | 62.3±23.1 | 69.2±22.3 | <.001† |
| Hypertension, no.<br>(%) | 1021 (90.8) | 475 (91.3) | 338 (90.1) | 208 (90.8) | .83‡ |
| Diabetes mellitus,<br>no. (%) | 394 (35.1) | 181 (34.8) | 137 (36.5) | 76 (33.2) | .70‡ |
| Current or recent<br>smoker, no. (%) | 37 (3.3) | 17 (3.3) | 10 (2.7) | 10 (4.4) | .53‡ |
| On dialysis, no. (%) | 24 (2.1) | 12 (2.3) | 6 (1.6) | 6 (2.6) | .69‡ |
| Immunocompromise<br>d | 137 (12.2) | 59 (11.3) | 41 (10.9) | 37 (16.2) | .12‡ |
| Peripheral vascular disease, no. (%) | 610 (54.3) | 298 (57.3) | 189 (50.4) | 123 (53.7) | .12‡ |
| Atrial fibrillation/flutter, no. (%) | 462 (41.1) | 222 (42.7) | 156 (41.6) | 84 (36.7) | .30‡ |
| Prior endocarditis, no. (%) | 13 (1.2) | 9 (1.7) | 2 (0.5) | 2 (0.9) | .25‡ |
| Prior myocardial infarction, no. (%) | 251 (22.3) | 114 (21.9) | 78 (20.8) | 59 (25.8) | .35‡ |
| Prior stroke, no. (%) | 97 (8.6) | 46 (8.8) | 28 (7.5) | 23 (10.0) | .53‡ |
| Prior transient ischemic attack, no. (%) | 89 (7.9) | 39 (7.5) | 28 (7.5) | 22 (9.6) | .59‡ |
| Prior CEA or CAS, no. (%) | 48 (4.3) | 18 (3.5) | 22 (5.9) | 8 (3.5) | .18‡ |
| Chronic pulmonary disease, no. (%) | 538 (47.9) | 226 (43.5) | 170 (45.3) | 102 (44.5) | .12‡ |
| Moderate, no. (%) | 186 (34.6) | 91 (40.3) | 64 (37.6) | 31 (30.4) | .41‡ |
| Severe, no. (%) | 114 (21.1) | 54 (23.9) | 38 (22.4) | 22 (21.6) | .41‡ |
| On home oxygen, no. (%) | 64 (5.7) | 29 (5.6) | 22 (5.9) | 13 (5.7) | .99‡ |
| Prior cardiac surgery, no. (%) | 326 (29.0) | 173 (33.3) | 93 (24.8) | 60 (26.2) | .02‡ |
| STS risk score,<br>median (IQR), % | 4.9 (3.1-8.0) | 5.0 (3.2-8.0) | 5.0 (3.0-8.0) | 4.9 (3.0-7.9) | .40† |
| Prior aortic valve<br>procedure, no. (%) | 133 (11.8) | 77 (14.8) | 35 (9.3) | 21 (9.1) | .02‡ |
| Balloon<br>valvuloplasty, no.<br>(%) | 29 (21.8) | 12 (15.6) | 9 (25.7) | 8 (38.1) | .63‡ |
| Surgical repair or<br>replacement, no. (%) | 97 (72.9) | 62 (80.5) | 22 (62.9) | 13 (61.9) | .001‡ |
| TAVR, no. (%) | 7 (5.3) | 3 (3.9) | 4 (11.4) | 0 | .33‡ |
| Prior non-aortic<br>valve procedure, no.<br>(%) | 40 (3.6) | 21 (4.0) | 14 (3.7) | 5 (2.2) | .44‡ |
| Prior CABG, no.<br>(%) | 250 (22.2) | 123 (23.7) | 75 (20.0) | 52 (22.7) | .42‡ |
| Prior PCI, no. (%) | 437 (38.9) | 202 (38.8) | 156 (41.6) | 79 (34.5) | .22‡ |
| Implantable<br>cardioverter<br>defibrillator, no. (%) | 25 (2.2) | 8 (1.5) | 9 (2.4) | 8 (3.5) | .23‡ |
| Permanent<br>pacemaker, no. (%) | 129 (11.5) | 64 (12.3) | 45 (12.0) | 20 (8.7) | .34‡ |
| Primary TAVR<br>procedure indication |  |  |  |  | .01‡ |
| Aortic stenosis,<br>no. (%) | 1028 (91.5) | 459 (88.3) | 350 (93.3) | 219 (95.6) |  |
| Aortic<br>regurgitation, no.<br>(%) | 18 (1.6) | 9 (1.7) | 8 (2.1) | 1 (0.4) |  |
| NYHA class in the<br>last 2 weeks |  |  |  |  | <.001‡ |
| III, no. (%) | 660 (58.7) | 326 (62.7) | 211 (56.3) | 123 (53.7) |  |
| IV, no. (%) | 93 (8.3) | 63 (12.1) | 20 (5.3) | 10 (4.4) |  |
| Creatinine, median<br>(IQR), mg/dL | 1.1 (0.9-1.4) | 1.1 (0.9-1.4) | 1.1 (0.9-1.4) | 1.1 (0.9-1.4) | .72† |
| Hemoglobin, g/dL | 12.4±1.9 | 12.2±1.9 | 12.5±1.9 | 12.5±1.7 | .05† |
| Platelet count,<br>median (IQR),<br>x10<sup>3</sup>/μl | 198.0 (161.0-244.0) | 200.0 (164.0-248.0) | 196.0 (162.0-244.3) | 195.0 (152.0-241.0) | .42† |
| Left ventricular<br>ejection fraction,<br>median (IQR), % | 61.0 (53.0-66.0) | 61.0 (50.0-65.0) | 62.0 (55.0-66.0) | 61.0 (55.0-65.5) | .03† |
| Bicuspid aortic<br>valve morphology,<br>no. (%) | 19 (1.7) | 12 (2.3) | 6 (1.6) | 1 (0.4) | .27‡ |
| Degenerative aortic valve disease, no. (%) | 1063 (94.6) | 492 (94.6) | 349 (93.1) | 222 () | .17‡ |
| Non-aortic valvulopathy severity ≥ moderate, no. (%) | 339 (30.2) | 176 (33.8) | 101 (26.9) | 62 (27.1) | .05‡ |
| Aortic valve area, median (IQR), cm<sup>2</sup> | 0.9 (0.7-1.0) | 0.9 (0.7-1.0) | 0.9 (0.7-1.0) | 0.9 (0.7-0.9) | .30† |
| Aortic valve mean gradient, mmHg | 42.7±13.2 | 41.9±13.6 | 43.3±12.5 | 43.7±13.6 | .16† |
| Aortic valve peak velocity, m/s | 4.1±0.7 | 4.1±0.7 | 4.1±0.6 | 4.2±0.7 | .21† |
| ≥1 diseased coronary artery on diagnostic catheterization in the last 12 months, no. (%) | 1108 (98.6) | 515 (99.0) | 368 (98.1) | 225 (98.3) | .51‡ |

\* Plus-minus values represent means ± SD. For skewed distributions, the medians and the interquartile ranges were provided. Group-1: significant improvement in KCCQ-12 score ( ≥ +19 points), group-2: modest improvement in KCCQ-12 (1-19 points), and group-3: no improvement in KCCQ-12 score ( ≤ 0 points).
† The *P* value was calculated using a one-way analysis of variance (ANOVA) or Kruskal-Wallis test.
‡ The *P* value was calculated using a chi-square or Fisher's exact test.
§ The BMI is equal to the weight in kilograms divided by the square of the height in meters.
Abbreviations: BMI, body mass index; IQR, interquartile range; KCCQ-12, Kansas City Cardiomyopathy Questionnaire 12; CEA, carotid endarterectomy; CAS, carotid artery stenting; STS, Society of Thoracic Surgeons; CABG, coronary artery bypass grafting; PCI, percutaneous coronary intervention; TAVR, transcatheter aortic valve replacement; NYHA, New York Heart Association.

At the early follow-up visit, 46.3% (n= 520) of patients had a significant improvement in their KCCQ-12 score, while 33.4% (n= 375) and 20.4% (n = 229) had modest and no improvement, respectively. The majority of the baseline characteristics were similar across the 3 groups; however, patients in group-1 had a higher median BMI (29.7; IQR, 25.7-34.1), a lower median LVEF (61.0%; IQR, 50.0-65.0), and were more likely to have had a prior cardiac surgery (33.3%), a prior AV procedure (14.8%), and an NYHA class of III or IV (74.8%). Additionally, patients in group-1 had a significantly lower mean KCCQ-12 score at baseline (40.7±18.7 points) compared to groups 2 and 3 (62.3±23.1 and 69.2±22.3 points, respectively). Patients in group-3 were significantly more likely to have AS as the primary indication for undergoing TAVR (95.6%) compared to patients in the other 2 groups.

### Procedural and in-hospital data

Overall implant success was 98.9% and was not significantly different between the cohorts (**Table 2**). Patients in group-3 had a significantly higher pacemaker requirement after the procedure relative to groups 1 and 2 (27.1 vs 17.5 vs 17.3, respectively; *P*=.004). Patients in group-1 were less likely to undergo TAVR electively (86.2%) and require cardiopulmonary bypass during the procedure (0.2%) but were more likely to have a valve-in-valve procedure (13.5%). The remaining procedural and in-hospital characteristics were comparable amongst the groups.

**Table 2.**
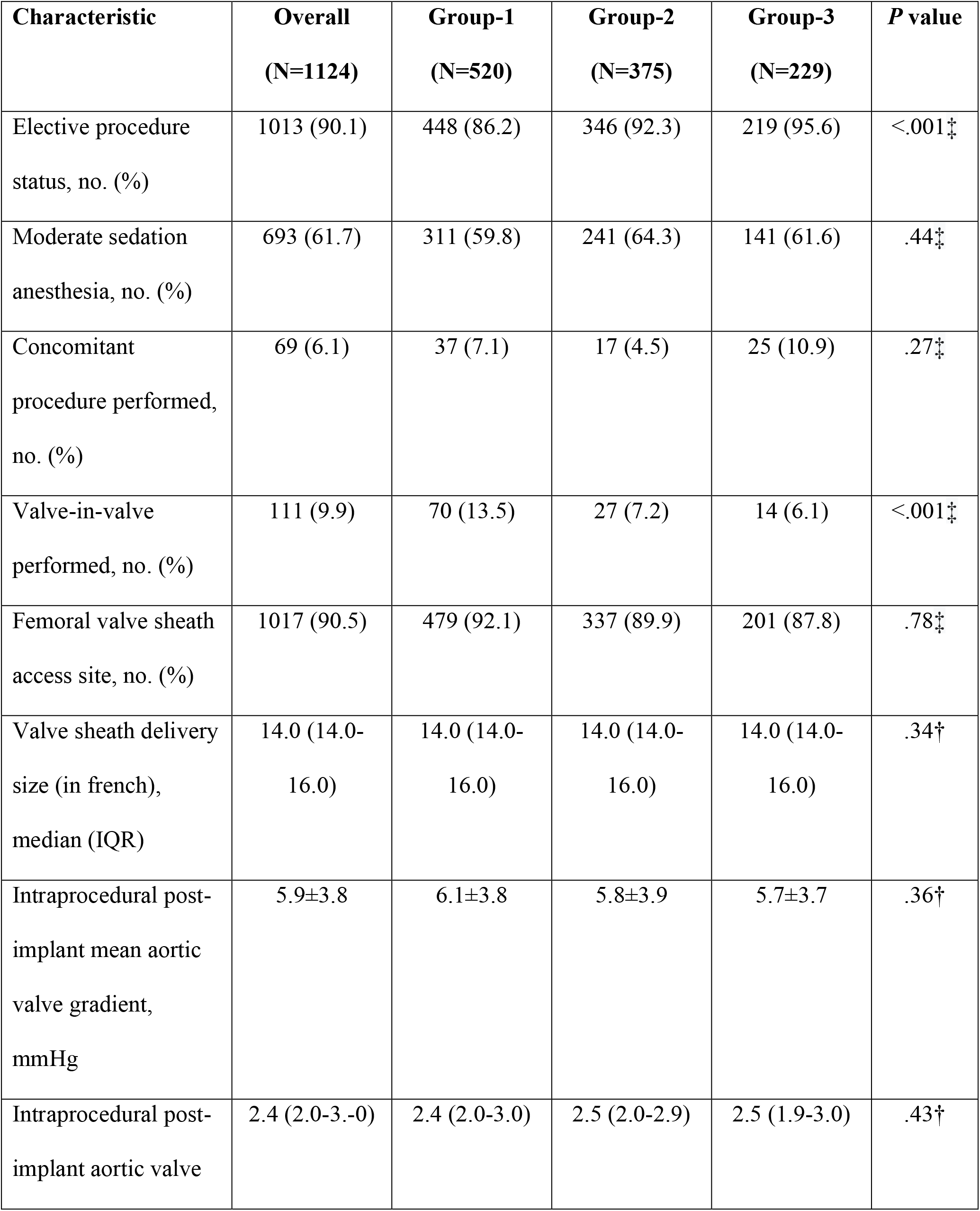
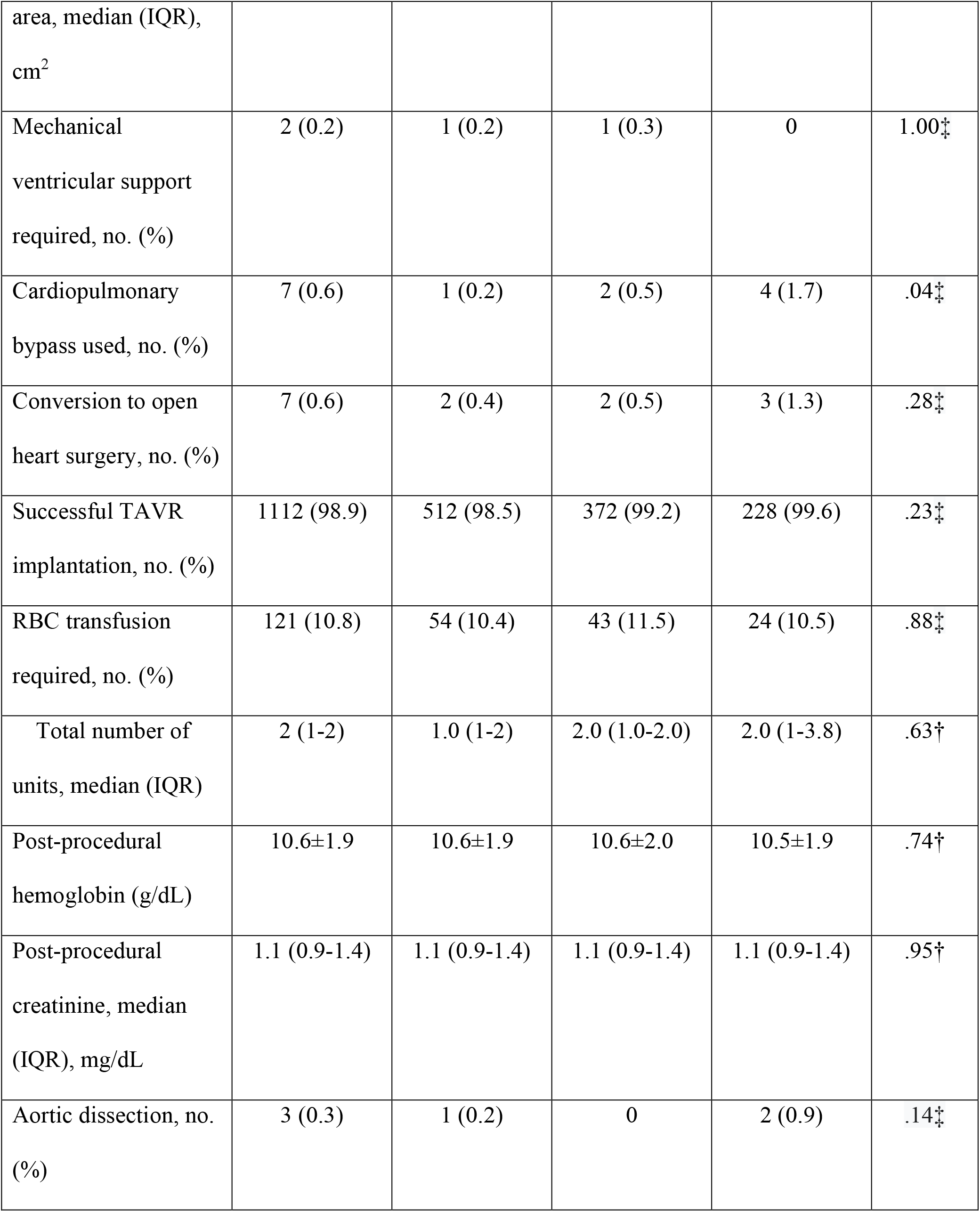
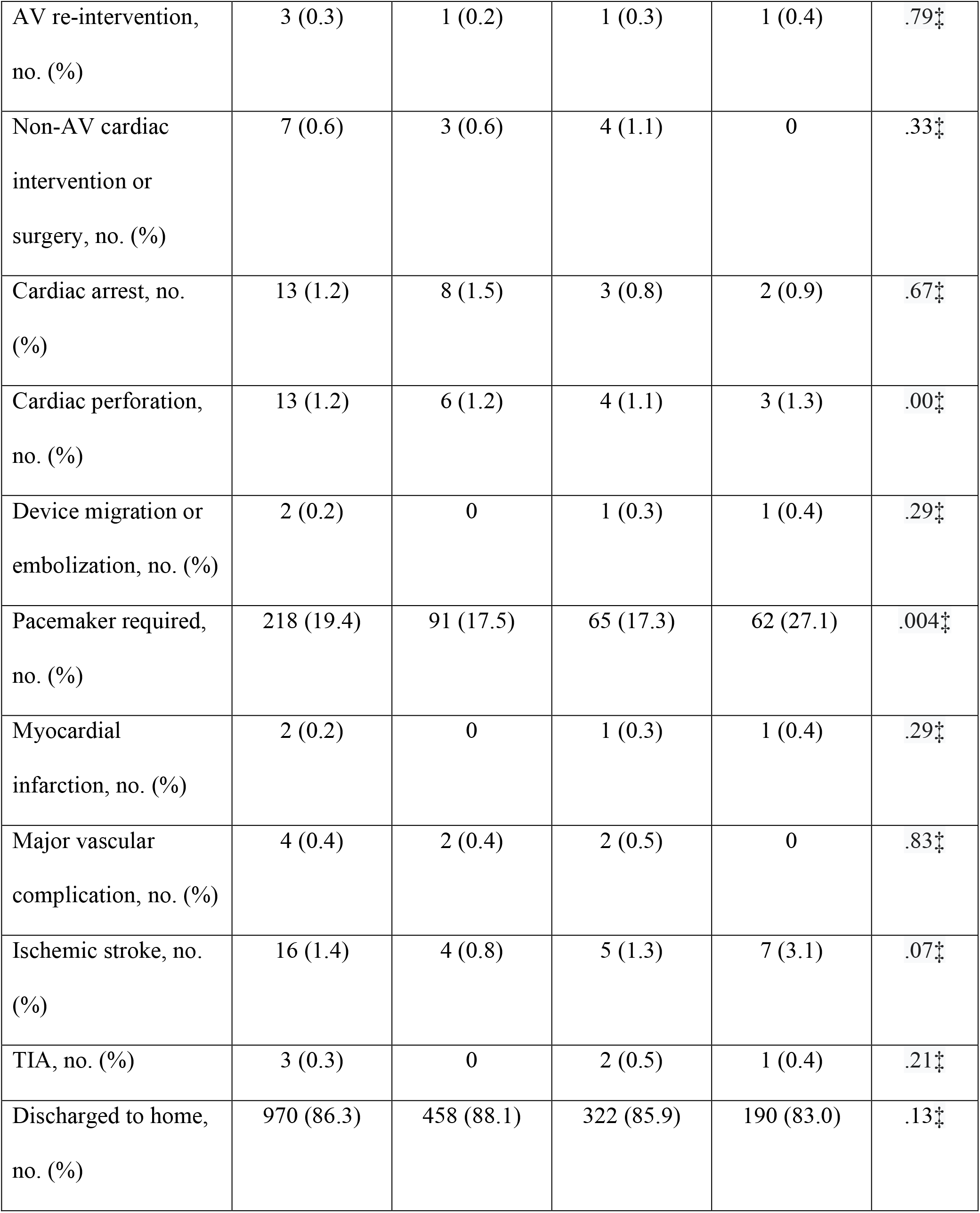
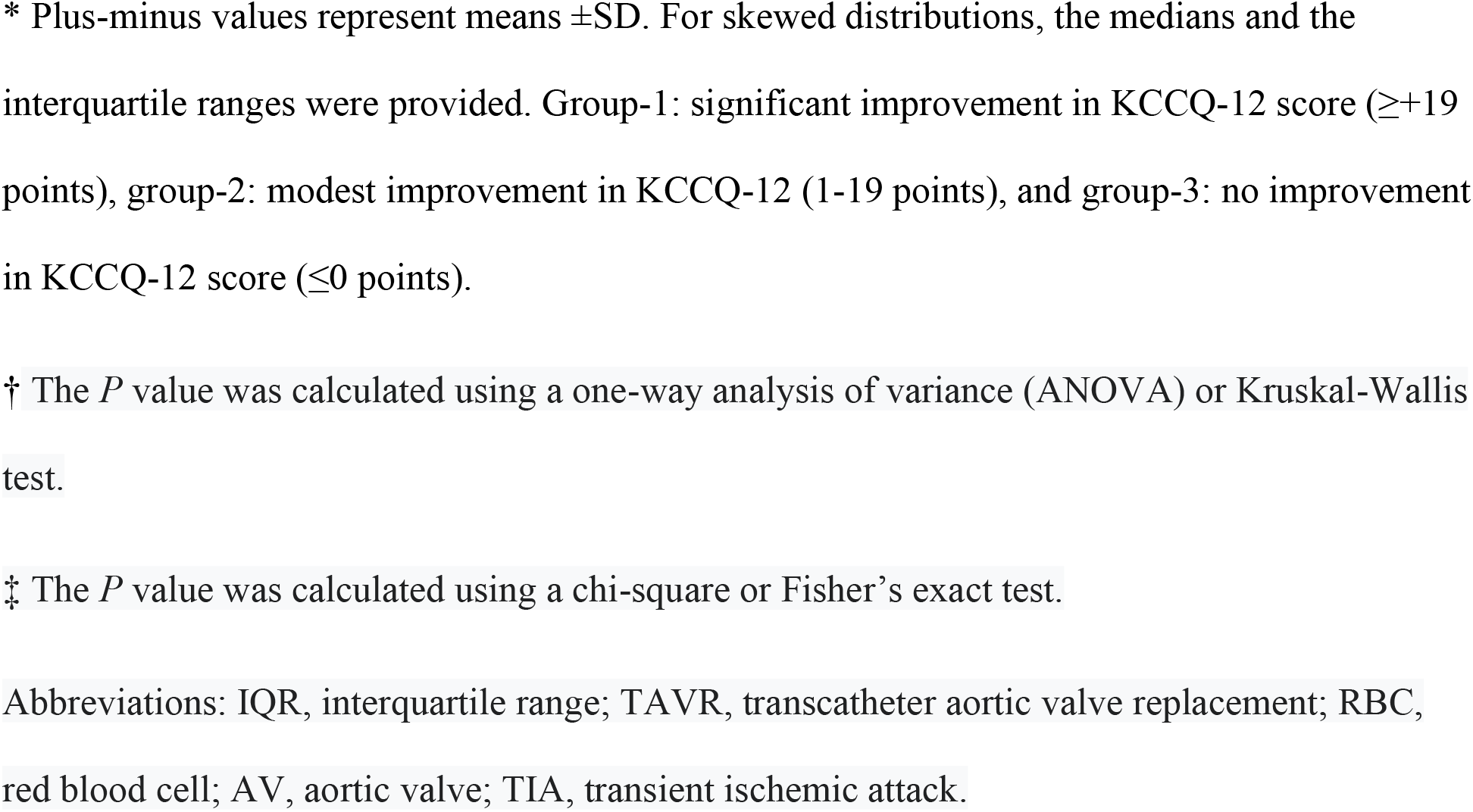
Overall and Comparison of Procedural and In-hospital Characteristics*.

### Early follow-up visit

At 45 day follow up, the overall mean (±SD) KCCQ-12 score was 72.8±22.3 points, with a mean increase of 19±24.0 points from baseline (**Table 3)**. The mean KCCQ-12 score was significantly different between the three cohorts after TAVR; patients in group-1 had a mean score of 80.2±15.5 while patients in groups 2 and 3 had a mean score of 72.4±22.6 and 58.3±10.8, respectively (*P*<.001). The prevalence of NYHA classes III and IV decreased from 67% to 5.1% after TAVR overall, with patients in group-1 having the lowest prevalence compared to the groups 2 and 3 (1.5% vs 5.1% vs 13.1, respectively; *P*<.001). However, patients in group-1 tended to have a significantly higher AV mean gradient (12.6±5.5 mmHg) and lower reduction in their AV mean gradient after TAVR (−29.4±14.0), relative to the other 2 groups.

**Table 3.** Overall and Comparison of Early Follow-up Visit Characteristics*.

| Characteristic | Overall<br>(N=1124) | Group-1<br>(N=520) | Group-2<br>(N=375) | Group-3<br>(N=229) | P value |
| --- | --- | --- | --- | --- | --- |
| NYHA class in the last 2 weeks |  |  |  |  | <.001<sup>‡</sup> |
| III, no. (%) | 48 (4.3) | 8 (1.5) | 19 (5.1) | 21 (9.2) |  |
| IV, no. (%) | 9 (0.8) | 0 | 0 | 9 (3.9) |  |
| KCCQ-12 score | 72.8±22.3 | 80.2±15.5 | 72.4±22.6 | 58.3±10.8 | <.001<sup>†</sup> |
| Change in KCCQ-12 score from baseline | 19.0±24.0 | 39.5±15.5 | 9.5±5.4 | -12.3±12.3 | <.001<sup>†</sup> |
| Hemoglobin, g/dL | 11.9±1.8 | 12.0±1.8 | 11.9±1.8 | 11.7±1.8 | .05<sup>†</sup> |
| Creatinine, median (IQR), mg/dL | 1.1 (0.9-1.4) | 1.1 (0.9-1.4) | 1.1 (0.9-1.4) | 1.1 (0.9-1.3) | .34<sup>†</sup> |
| Left ventricular ejection fraction, median (IQR), % | 61.0 (54.0-65.0) | 60.0 (52.0-65.0) | 61.0 (55.0-65.0) | 61.0 (55.0-65.0) | .25<sup>†</sup> |
| Aortic valve mean gradient, mmHg | 12.4±5.7 | 12.6±5.5 | 12.5±6.3 | 11.5±5.0 | .04<sup>†</sup> |
| Change in aortic valve mean gradient from baseline, mmHg | -30.5±13.6 | -29.4±14.0 | -30.9±13.0 | -32.3±13.7 | .03<sup>†</sup> |
| Aortic valve regurgitation severity ≥moderate | 60 (5.3) | 24 (2.1) | 14 (3.7) | 22 (9.6) | .01‡ |

\* Plus-minus values represent means ± SD. For skewed distributions, the medians and the interquartile ranges were provided. Group-1: significant improvement in KCCQ-12 score ( $\geq +19$ points), group-2: modest improvement in KCCQ-12 (1-19 points), and group-3: no improvement in KCCQ-12 score ( $\leq 0$ points).
† The *P* value was calculated using a one-way analysis of variance (ANOVA) or Kruskal-Wallis test.
‡ The *P* value was calculated using a chi-square or Fisher's exact test.
Abbreviations: NYHA, New York Heart Association; KCCQ-12, Kansas City Cardiomyopathy Questionnaire 12; IQR, interquartile range.

### Long-term survival and the prognostic value of the KCCQ-12 score

Over a median follow-up of 3.0 years, the Kaplan-Meier survival analysis revealed an overall survival time of 5.4 years (95% CI, 5.0-5.7) for the entire study population. A significantly higher median survival time of 5.7 years (95% CI, 5.2-6.2) was observed for patients in group-1 compared to 5.1 years (95% CI, 4.5-5.8) and 4.1 years (95% CI, 3.4-4.9) for patients in group 2 and 3, respectively (*P* by log-rank <.001) as shown in Figure 1A. After adjusting for multiple potential confounders, a Cox-proportional hazards analysis showed an association between modest improvement in KCCQ-12 and all-cause mortality (adjusted hazard ratio (HR), 1.64; 95% CI, 1.28-2.10) or no improvement (adjusted HR 2.44; 95%CI, 1.84-3.24) (**Figures 1B and 2**). Additionally, preprocedural KCCQ-12 score alone was significantly correlated with long-term mortality, with an 8.0% reduction in mortality risk being observed for every 5-point increase in the score (*P*<.001). Other independent predictors of long-term mortality included preprocedural atrial fibrillation, immunocompromised status, hemodialysis, chronic pulmonary disease, hypertension, and STS (**Figure 2**).

**Figure 1.**
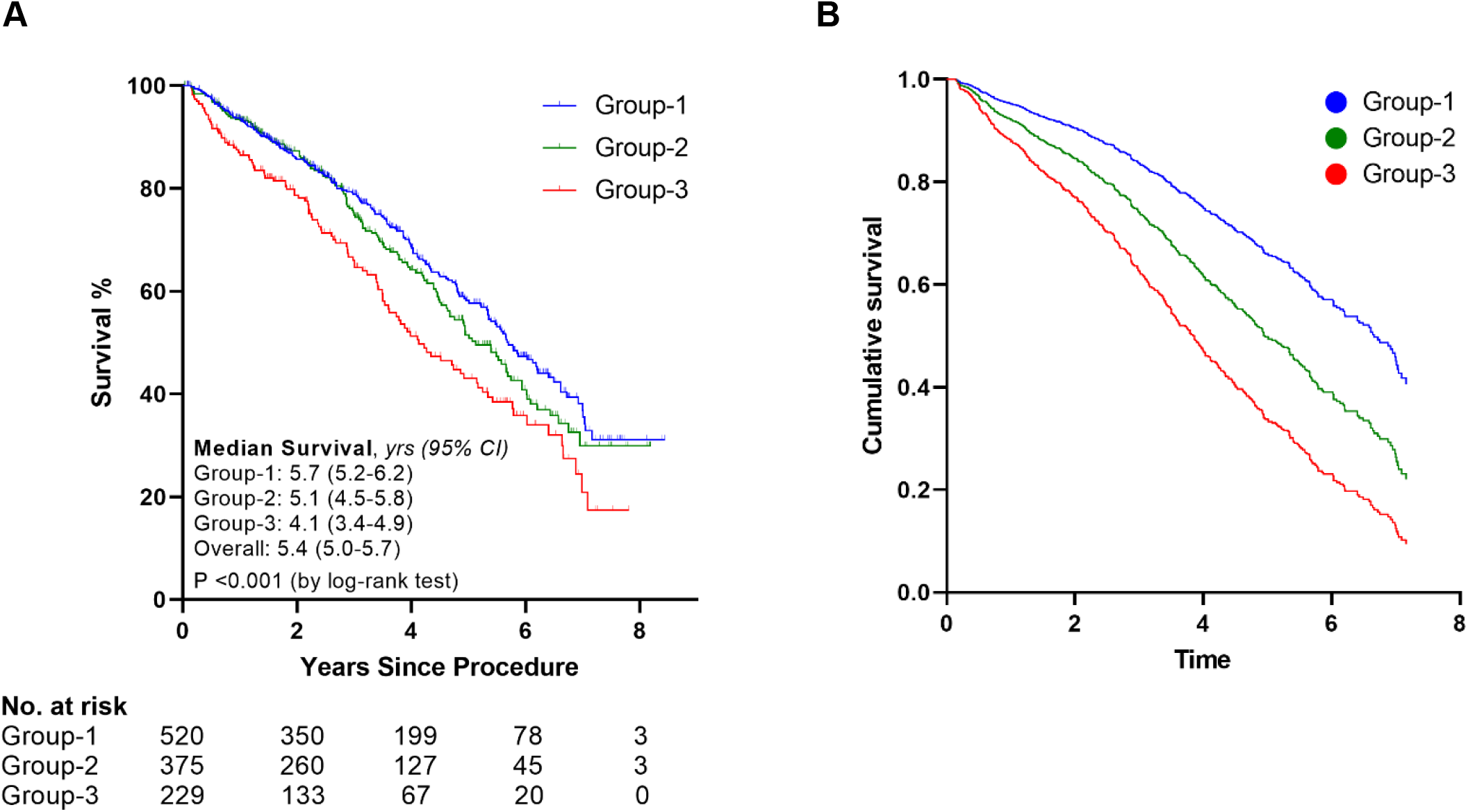
Long-term Survival Based on Improvement in KCCQ-12 Score at the Early Follow-up Visit*. **Panel A** represents the Kaplan-Meier curves for groups 1-3 along with the number at risk (at 2-year intervals) and the median survival time after the procedure. **Panel B** depicts the Cox-proportional hazards regression curves for groups 1-3 after adjusting for the following variables: age, atrial fibrillation/flutter, baseline aortic valve mean gradient, baseline left ventricular ejection fraction, baseline NYHA class, baseline KCCQ-12 score, BMI, chronic pulmonary disease, diabetes mellitus, hemodialysis, hypertension, immunocompromised status, postprocedural pacemaker requirement, primary TAVR indication, prior myocardial infarction, prior cardiac surgery, prior stroke or transient ischemic attack (TIA), sex, smoking status, and STS risk score. * Group-1: significant improvement in KCCQ-12 score (≥ +19 points), group-2: modest improvement in KCCQ-12 (1-19 points), and group-3: no improvement in KCCQ-12 score (≤ 0 points).

**Figure 2.**
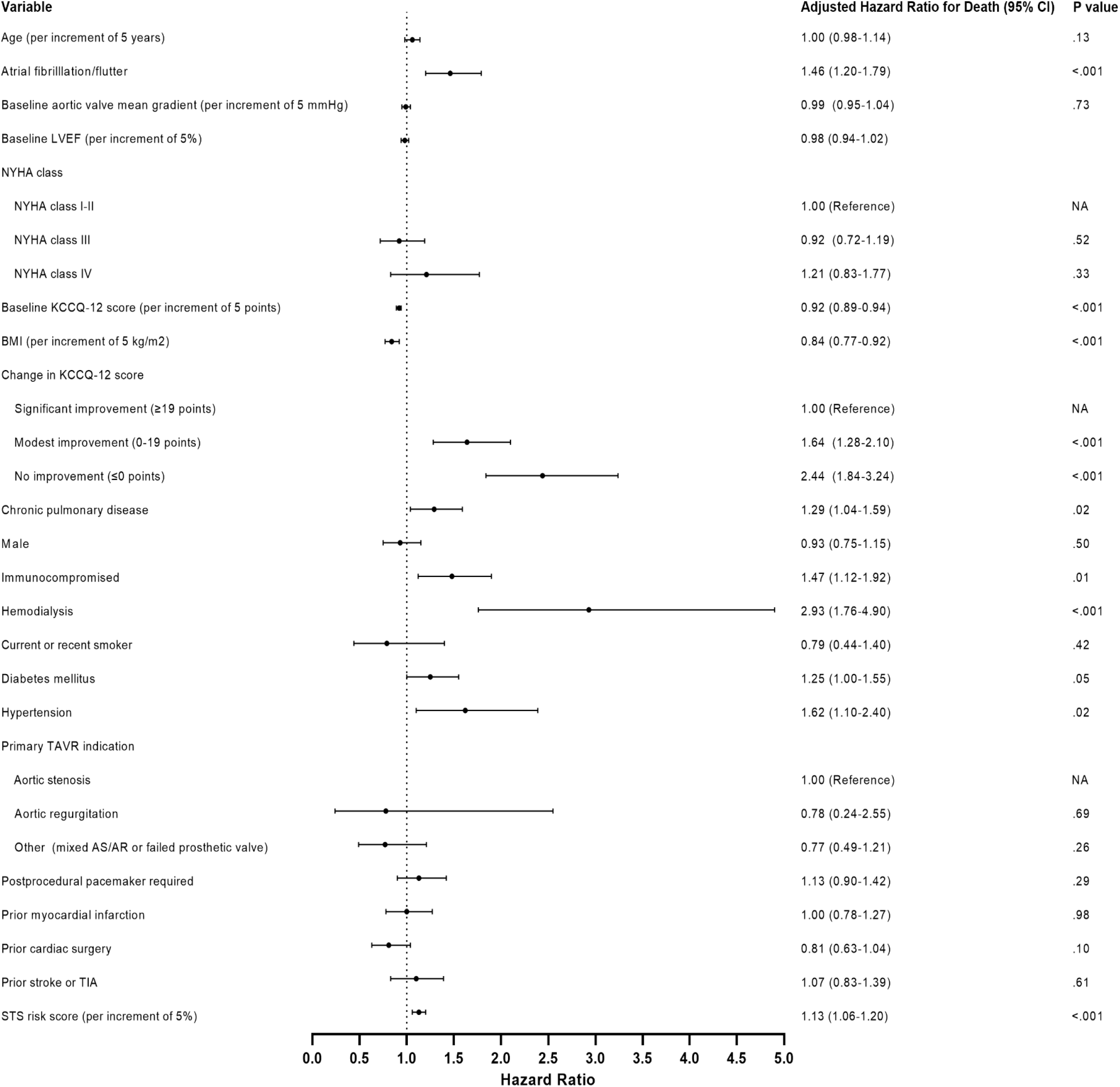
Association of Different Variables with Long-term Mortality in TAVR*. The Forest plot depicts the adjusted hazard ratios for death in TAVR patients after adjusting for multiple variables as measured by a multivariable cox-proportional hazards regression model. Abbreviations: LVEF, left ventricular ejection fraction; NYHA, New York Heart Association; NA, not applicable; KCCQ, Kansas City Cardiomyopathy Questionnaire; TAVR, transcatheter aortic valve replacement; AS/AR, Aortic stenosis/Aortic regurgitation; TIA, transient ischemic attack; STS, Society of Thoracic Surgeons. *Body mass index, baseline left ventricular ejection fraction, baseline KCCQ-12 score, and STS risk score were evaluated as continuous variables. Adjusted hazard ratios for those variables were depicted per 5-unit increments.

## Discussion

Our study documents 2 key findings: first, it demonstrates that changes in the KCCQ-12 score after TAVR are a significant, independent prognostic factor for long-term survival, with modest and no improvement in KCCQ-12 score after the procedure being associated with a considerably higher long-term mortality risk. Second, lower preprocedural KCCQ-12 score was associated with lower long-term survival after TAVR. Those findings are deserving of further elucidation.

The ability of patient-reported outcome measures (PROMs) to yield critical prognostic information has previously been validated for various medical conditions such as AS, HF, and cancer.^6,10,14^ In patients with HF with reduced (HFrEF) and preserved (HFpEF) ejection fraction, KCCQ has consistently demonstrated excellent prognostic qualities with regard to all-cause mortality, cardiovascular death, and hospitalization.^3–5,15^ The KCCQ has also been shown to be a reliable predictor of 1-year resource use and costs in patients with HFrEF after a myocardial infarction.^16^ When compared with the NYHA classification system, the 6-minute walk test, and the Minnesota Living with Heart Failure Questionnaire (MLHFQ), KCCQ has demonstrated superior sensitivity to clinically meaningful changes in patients with HF while outperforming both, the MLHFQ and NYHA class, in predicting outcomes related to mortality and hospitalization.^15,17^

The utility of KCCQ in risk stratification of patients undergoing TAVR has been explored previously. In a study by Arnold et al. (2012), patients with severe, symptomatic AS who were randomized to either medical management, SAVR, or TAVR had quality of life assessment with the KCCQ tool before and after the intervention.^7^ Overall, different domains of the KCCQ were found to be a valid measure of change in symptoms, functional status, and quality of life when compared with other instruments that captured similar information. The study also evaluated the association between baseline KCCQ scores and 1-year mortality in patients who were randomized to medical therapy only. In those patients, a low baseline KCCQ was found to be strongly associated with an increased risk of mortality. In a subsequent study by Arnold et al, patients who had poor (25 to 49 points), or very poor (<25 points) health status had much higher mortality than those with good baseline health status (KCCQ score ≥ 75 points) (adjusted HR, 2.00, and adjusted HR, 1.54, respectively).^11^ However, these studies only included baseline KCCQ and did not examine the association of its changes after the procedure with clinical outcomes.

Recently, Hejjaji et al. attempted to bridge this gap by examining the prognostic importance of the pre-and post-procedural KCCQ-12 (assessed at 30 days), and the change in KCCQ-12 (from baseline to 30 days post-TAVR) in predicting 1-year mortality and HF hospitalization after TAVR.^10^ In the risk-adjusted model, the change in KCCQ score at 30 days was significantly associated with 1-year mortality risk, with a larger change in score being more strongly associated with a lower mortality (adjusted HR (per 5 points), 0.93; 95% CI, 0.92-0.94). However, these data were limited to 1 year follow up and was hindered by the heterogeneity of KCCQ performance and reporting across site. Our study corroborates those findings with a more extended follow-up to an average of 3 years. These findings have some practical implications. They illustrate the utility of serial measurements of KCCQ-12 in the management of patients with AS following TAVR. Patients with limited changes in KCCQ-12 could benefit from a targeted more intensive follow up protocol to address their residual risk. The changes in KCCQ-12 after TAVR can further be used to study the utility/futility and the cost-effectiveness of TAVR and may be other interventions in the future.

Another interesting observation in our study was the inverse association between baseline BMI and long-term survival in TAVR patients, a finding that is in line with prior research documenting this phenomenon.^18,19^ Our study further identifies patients with a baseline BMI <23 to be at a notably higher risk of long-term mortality after TAVR, which suggests that this could be a reasonable marker of frailty in patients undergoing the procedure. A similar association was also observed in Winter et al.’s (2014) meta-analysis of the general population above the age of 65 years.^20^

Finally, the results of our study must be interpreted with caution considering its several limitations. First, this is a single-center study that was performed at an academic, tertiary center; thus, despite its large sample size, generalizability may still be limited. Second, although interest and emphasis on PROMs has risen through the years, their incorporation in practice remains challenging and inconsistent. Therefore, we were unable to include all patients who underwent TAVR during that time due to missing KCCQ-12 data, which may also impact the generalizability of our results. Third, although we attempted to adjust for multiple potential confounders and baseline differences between the groups, we cannot exclude the possibility of residual confounding attributable to variables that were not captured by the ACC/STS TVT registry.

## Conclusion

Patients with a lower baseline KCCQ-12 score and those who experience modest or no-improvement in their KCCQ-12 score after TAVR have a substantially higher long-term mortality risk. The KCCQ-12 is a cost-effective, efficient tool that can help identify patients at increased risk of death after TAVR, thereby assisting clinicians and patients in making more informed decisions pre-and post-procedurally.

## Data Availability

De-identified data can be shared by the corresponding author upon request.

## Nonstandard Abbreviations and Acronyms

TAVR: Transcatheter aortic valve replacement
AS: Aortic stenosis
KCCQ: Kansas City Cardiomyopathy Questionnaire
STS: Society of Thoracic Surgeons

## Acknowledgements

None.

## Sources of Funding

None.

## Disclosures

None.

## Notes

**Disclosures:** All authors have no relevant conflicts to disclose.

### Competing Interest Statement

The authors have declared no competing interest.

### Author Declarations

Mayo Clinic - Rochester's IRB.

